# Comparative Risk of the Onset of Atrial Fibrillation after Icosapent Ethyl versus Omega-3 –Acid-Ethyl-Esters Adjuvant to Statins

**DOI:** 10.1101/2024.09.16.24313779

**Authors:** Jyotirmoy Sarker, Michael Kim, Samantha Patton, Przemysław B. Radwański, Mark A. Munger, Kibum Kim

**Affiliations:** Department of Pharmacy Systems, Outcomes and Policy, University of Illinois Chicago, Chicago, Illinois; Department of Pharmacotherapy, University of Utah Health, Salt Lake City, Utah; 2Department of Physiology and Cell Biology, The Ohio State University, Columbus, Ohio, USA; The Frick Center for Heart Failure and Arrhythmia, Dorothy M. Davis Heart and Lung Research Institute, College of Medicine, The Ohio State University Wexner Medical Center, Columbus, OH, US; Division of Pharmacology and Pharmaceutics, College of Pharmacy, The Ohio State University, Columbus, OH, US; Department of Internal Medicine, University of Utah Health, Salt Lake City, Utah

**Keywords:** Cohort Study, Icopasent Ethyl, Omega-3 Fatty Acids, Atrial Fibrillation

## Abstract

**Background:** Icosapent ethyl (ICP), an ethyl ester of eicosapentaenoic acid (EPA), and omega-3 acid ethyl esters (DHA/EPA), comprised of ethyl esters of EPA and doxosahexaenoic acid (DHA), are approved as adjunctive therapy to statins for reducing adverse cardiovascular events (CV) in patients with CV risks. However, there are concerns regarding a potential association between ICP and atrial fibrillation (AF). This study evaluated the incidence of AF onset between ICP and DHA/EPA when used as adjuvant therapy with statins.

**Methods and Results:** This retrospective study utilized administrative healthcare claims to analyze adult AF-naïve patients from one year preceding their first prescription for ICP or DHA/EPA. These patients were followed for two years, spanning from2013-2021. AF incidence was assessed during active treatment with either ICP or DHA/EPA as adjunct statin therapy. A propensity score (PS) matched cohort controlled for baseline characteristics and the effect of calendar year on the use of ICP or DHA/EPA. The cumulative incidence of AF was estimated using a product-limit estimator and compared between groups using a Cox proportional hazards regression model. The PS-matched cohort included 17,638 participants with a mean age 56 years, predominantly male (65.7% ICP vs. 64.5% DHA/EPA). Over two years, the cumulative incidence of AF from ICP and DHA/EPA was 5.32% and 3.99% respectively, resulting in a HR of 1.242 (95% CI: 1.061 to 1.455).

**Conclusions:** In adult AF-naïve patients, ICP, when compared to DHA/EPA in conjunction with statin therapy, was associated with a significantly higher significant risk of developing AF.

**RESEARCH PERSPECTIVE:** *What is New?:*

- Does icopasent ethyl (ICP), an ethyl ester of eicosapentaenoic acid (EPA) and omega-3 acid ethyl esters comprised of ethyl esters of EPA and doxosahexaenoic acid (DHA) in atrial fibrillation (AF)-naïve patients taking statins increase the incidence of AF?
- Over two years, the cumulative incidence of AF from ICP and DHA/EPA was 5.32% and 3.99% respectively, resulting in a HR of 1.242 (95% CI: 1.061 to 1.455).
- In adult AF-naïve patients, ICP, when compared to DHA/EPA in conjunction with statin therapy was associated with a higher significant risk of developing AF.

*What Question Should be Addressed Next?:*

- What should be considered as clinical and demographic factors in identifying patients at risk of atrial fibrillation prior to being prescribed ICP or DHA/EPA agents.
- Investigation into the underlying mechanism of the increase in atrial fibrillation with marine omega-3 ethyl esters should continue.
- Understanding AF outcomes from ICP or DHA/EPA use including AF burden, need for AF medical or electrophysiological interventions, and health-care total costs.

## INTRODUCTION

Dyslipidemia is a prevalent and significant health issue, contributing to a high burden of cardiovascular morbidity and mortality. Current guidelines recommend lipid-lowering therapy to manage dyslipidemia, with statin therapy being the cornerstone due to its role in lowering the risk of major adverse cardiovascular events (MACE). In patients with severe hypertriglyceridemia, icosapent ethyl (ICP) is indicated as an adjunct to maximally tolerated statin therapy to reduce the risk of MACE outcomes in adult patients with elevated TG levels (≥ 150mg/dL) and established CV disease or diabetes mellitus with two or more additional risk factors for CV disease.^1^ Besides ICP, which contains eicosapentaenoic acid (EPA), another FDA-approved prescription-available omega-3 acid ethyl esters contains EPA and docosahexaenoic acid (DHA).

ICP has been associated with an increased risk of AF requiring hospitalization. In the double-blind, placebo-controlled trial REDUCE-IT trial, it was reported that among 8,179 statin-treated patients with established CV disease or diabetes plus an additional risk factor for CV disease, 3.1% of patients treated with ICP experienced adjudicated AF or flutter requiring hospitalization for ≥ 24 hours, compared to 2.1% in the placebo arm [HR 1.5; 95% CI: 1.14-1.98].^2^ A recent post-hoc safety analysis of patients with or without prior AF before study randomization showed a significant 25% higher rate AF overall, with a 13% increase in patients with prior AF and a 26% increase in those without prior AF.^3^ These findings align with results observed after 2 years of 1.8g n-3 omega-3 fatty acid supplementation (i.e. DHA/EPA), which showed a significant increase in new AF events.^4^ Given that ICP has a higher concentration of EPA alone, there may be differences in the risk of AF onset compared to DHA/EPA.

This study aimed to address this gap by evaluating the onset of AF in patients naïve to AF who are initiating treatment with ICP compared to DHA/EPA, while on baseline statin therapy in a large, real-world dataset.

## METHODS

### Study design and population

We conducted a retrospective cohort study using an administrative healthcare claims database. Patient-level de-identified claims were abstracted from the Merative MarketScan Commercial Claims and Medicare Supplemental Databases that includes patients covered by employment-based insurance plans in the United States. The data source covered medical and outpatient pharmacy claims from January 2013 to December 2021. Direct patient identifiers were removed before investigators had access to the data. The University of Illinois Chicago Institutional Review Board deemed the use of the database for this study exempt.

### Eligibility Criteria and Patient Characteristics

A retrospective cohort study with an active comparator new user study design was performed. The target population was patients who are in need of lowering cardiovascular risk and are being treated with statin agents. Based on the outpatient drug dispensing records, we identified adult (≥ 18 years) patients who received either ICP (intervention) or DHA/EPA (control). The date of the first ICP or DHA/EPA dispensing was defined as the index date for this study. To be eligible, patients were required to have a dispensing of statin on the index date or to have an active supply of statin based on the most recent dispensing record prior to the index date.

The baseline period ensured that patients were previously naïve to intervention or control agents or AF and was 365-days before the index date. Eligible subjects were required to be enrolled in the study throughout the baseline period without any coverage gap. To identify AF, diagnosis codes ICD-9 427.31 and ICD-10 148.0, 148.1, 148.2, and 148.91 were used. We also excluded patients who had an AF encounter claim decided by the AF diagnosis codes within the first 30 days after ICP or DHA/EPA initiation, during which the risk carried over from the baseline period could outweigh the effect caused by the ICP or DHA/EPA use.

Patient characteristics were identified on the index date or during the baseline period. Demographic factors included age (by year and grouped), sex, region, insurance type, year of initiation, sedentary behavior, and substance abuse disorder. As potential confounders in the exposure-outcome association, we identified comorbid conditions including acute coronary syndrome, heart failure, type 1 diabetes mellitus, type 2 diabetes mellitus, hypertension, hypercholesterolemia, anxiety, depression, eating disorder, and hyperthyroidism. History of medication that might interact with or influence the prescription for fatty acid, statin therapy or CV risks included angiotensin II receptor blockers, angiotensin receptor-neprilysin inhibitors, beta blockers, calcium channel blockers, angiotensin-converting enzyme inhibitors (ACEi), angiotensin receptor blockers (ARBs), and glucoagon-like-peptide-1 receptor agonists (GLP-1ra) and sodium-glucose cotransporter-2 inhibitors (SGLT2i). Comorbidities were identified using International Classifications of Diseases version 9 and 10 (ICD-9 and ICD-10) diagnosis codes. We used national drug codes (NDCs)to screen the medication records.

To control the potential confounders in the measure of association, ICP participants were matched with DHA/EPA participants based on the propensity score (PS) and exact matching criteria. The PS of being assigned to ICP or DHA/EPA were calculated from a logistic regression model where baseline demographics and clinical characteristics deemed significantly different (*p*<0.05) between the ICP and DHA/EPA arms were utilized as independent variables. We applied nearest neighbor matching with a caliper of 0.1 range of pooled standard deviation of the logit of PS alongside with exact matching on the year of the index date in three groups (2013-15, 2016-2018, 2019-2021) by complete 1-by-1 matching. Standardized mean difference (SMD) of each variable was used as the balance diagnostics for the matched cohort.

Individuals were followed for up to two years. The primary outcome was the incidence of AF within two years of initiating ICP or DHA/EPA. A healthcare claim at either inpatient or outpatient setting decided by the diagnosis of AF at any position was considered for the outcome. Censoring from the follow-up occurred when the participant met any of the following criteria, whichever happened first: treatment discontinuation (defined as a gap of supply of more than 30 days), switching treatments, and disenrollment from the database before two years, or at the end of the study. Any discontinuation of the drugs of interest or statins was considered a treatment discontinuation. For example, if there was a continuous supply of ICP or DHA/EPA but a gap of more than 30 days of statin, the participant was discontinued and observations were censored.

### Statistical Analysis

We analyzed time to AF onset data using Kaplan-Meier estimates. The comparative outcome assessment was performed using Cox Proportional Hazards Regression model from which hazard ratio (HR) and 95% confidence intervals (95% CI) of the onset of AF for ICP and DHA/EPA were calculated. To control the residual confounders, we ran a series of multivariable regression models where each of the covariates showing *p*<0.05 were adjusted after matching.

## RESULTS

We identified a total of 292,961 participants who initiated treatment with ICP or DHA/EPA between January 1, 2013, and June 30, 2021. Of these, 67,285 participants were concurrently taking statins on the day of initiating ICP or DHA/EPA while meeting the baseline continuous enrollment criteria. After excluding 2,966 participants who had a diagnosis of AF prior to initiating ICP or DHA/EPA and removing patients who were censored or had AF event within the first 30 days, the potential participant pool included a total of 62,609 patients, consisting of 31,133 ICP participants and 31,476 DHA/EPA participants, eligible for matching [Figure 1].

**FIGURE 1:**
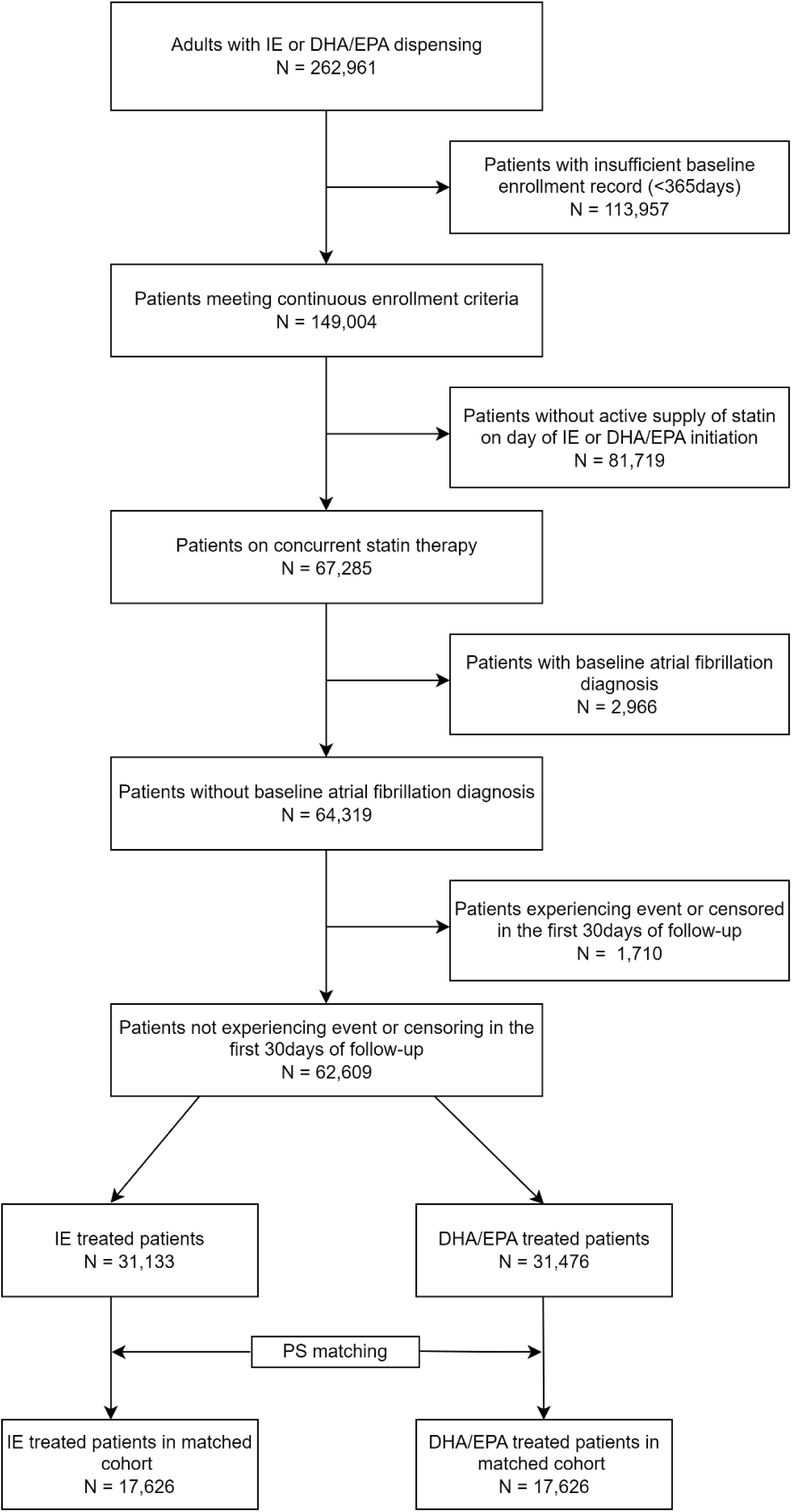
Cohort attrition diagram

**FIGURE 2:**
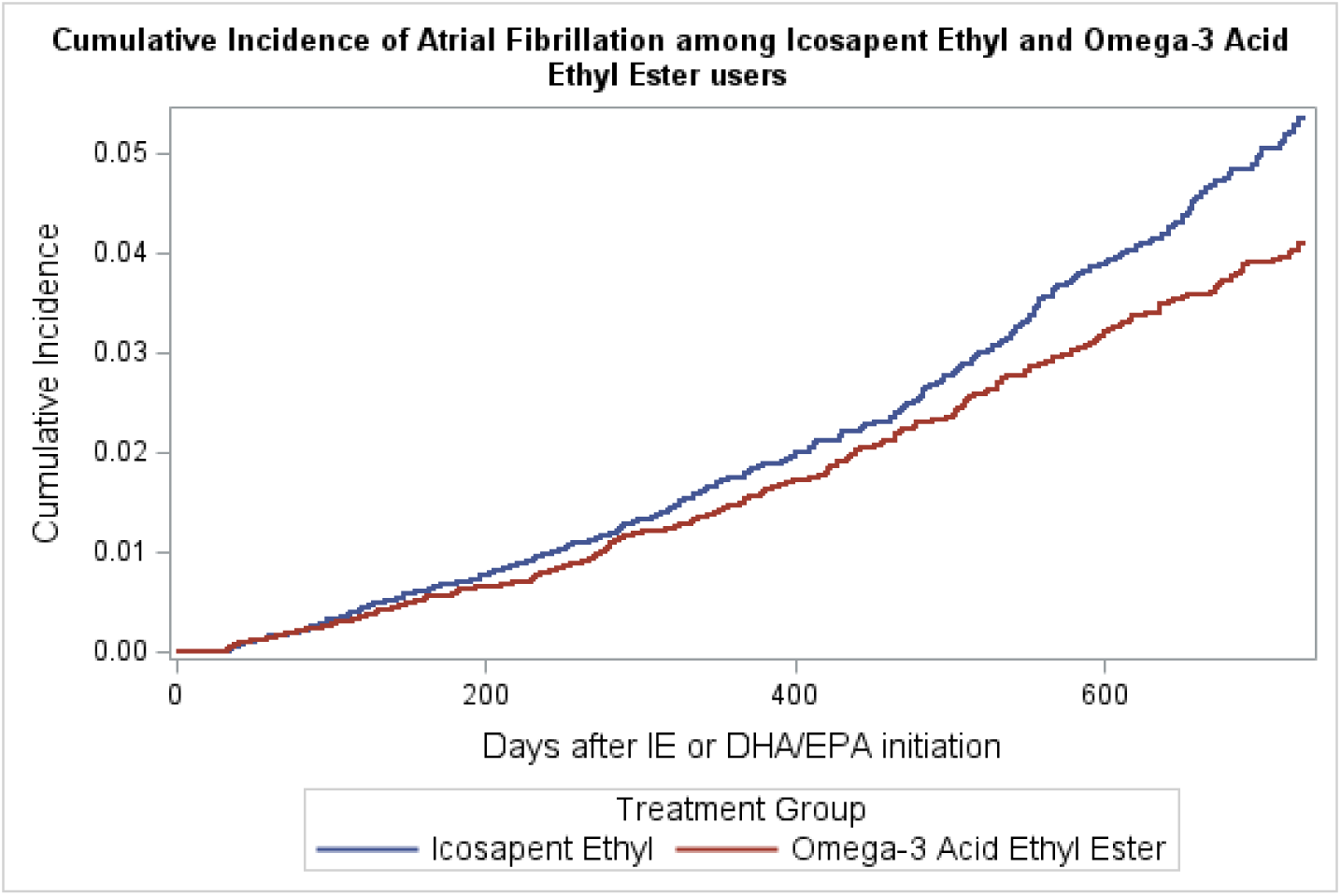
Cumulative incidence of atrial fibrillation among patients treated with icosapent ethyl and omega-3 acid ethyl ester.

The baseline characteristics of the two groups before PS matching are shown in Table 1. The age distribution differed between the groups, with a standardized mean difference (SMD) of 0.123. Specifically, a higher proportion of participants aged 65 and older initiated DHA/EPA (16.8%) compared to those initiating ICP (12.6%). Conversely, more participants aged 55-64 initiated ICP (42.9%) compared to DHA/EPA (39.8%). The proportion of male patients was higher in the ICP group (67.9%) compared to the DHA/EPA group (62.1%), with an SMD of 0.122. There was also a substantial difference in the distribution of participants across the study periods. More participants-initiated DHA/EPA in 2013-2015 (55.8%) compared to ICP (16.9%), whereas a higher proportion initiated ICP in the later years, 2019-2021 (56.9%) compared to DHA/EPA (14.7%). Insurance type also differed, with a larger proportion of participants on Medicare among the DHA/EPA participants (17.4%) compared to the ICP group (12.7%). Medication use patterns varied between the groups. For instance, baseline ARB use was more prevalent among ICP participants (30.1% vs. 26.0%, SMD = 0.09), while the proportion of patients on GLP-1ra/SGLT-2i use was markedly higher among the ICP group (24.0% vs. 10.9%, SMD = 0.35). Clinical conditions also differed. For example, hypertension (HTN) was more prevalent in the DHA/EPA group (25.2% vs. 13.4%, SMD = 0.301), as was type 2 diabetes mellitus (15.7% vs. 7.1%, SMD = 0.271).

**TABLE 1:** Baseline demographics and clinical characteristics.

|  |  | **IE** | **EPA/DHA** | **p** | **SMD** |
| --- | --- | --- | --- | --- | --- |
| **Characteristics** |  |  |  |  |  |
| **N** |  | 31133 | 31476 |  |  |
| **Age Group (%)** |  |  |  | <0.001 | 0.123 |
|  | 18-34 | 729 (2.3) | 787 (2.5) |  |  |
|  | 35-44 | 3,620 (11.6) | 3,668 (11.7) |  |  |
|  | 45-54 | 9,520 (30.6) | 9,215 (29.3) |  |  |
|  | 55-64 | 13,349 (42.9) | 12,525 (39.8) |  |  |
|  | 65+ | 3,915 (12.6) | 5,281 (16.8) |  |  |
| **Sex, Male (%)** |  | 21,151 (67.9) | 19,554 (62.1) | <0.001 | 0.122 |
| **Year (%)** |  |  |  | <0.001 | 1.091 |
|  | 2013-2015 | 5,274 (16.9) | 17,563 (55.8) |  |  |
|  | 2016-2018 | 8,135 (26.1) | 9,286 (29.5) |  |  |
|  | 2019-2021 | 17,724 (56.9) | 4,627 (14.7) |  |  |
| **Region (%)** |  |  |  | <0.001 | 0.207 |
|  | North Central | 4,059 (13.0) | 4,360 (13.9) |  |  |
|  | Northeast | 7,227 (23.2) | 7,456 (23.7) |  |  |
|  | South | 16,954 (54.5) | 15,210 (48.3) |  |  |
|  | Unknown | 71 (0.2) | 565 (1.8) |  |  |
|  | West | 2,822 (9.1) | 3,885 (12.3) |  |  |
| **Insurance type, Medicare (%)** |  | 3,966 (12.7) | 5,470 (17.4) | <0.001 | 0.13 |
| **Medication Use (%)** |  |  |  |  |  |
|  | ACEi | 10,161 (32.6) | 10,608 (33.7) | 0.005 | 0.023 |
|  | ARB | 9,358 (30.1) | 8,193 (26.0) | <0.001 | 0.09 |
|  | ARNI | 128 (0.4) | 32 (0.1) | <0.001 | 0.061 |
|  | Beta Blocker | 11,593 (37.2) | 10,630 (33.8) | <0.001 | 0.072 |
|  | Calcium Channel Blocker | 6,743 (21.7) | 6,653 (21.1) | 0.114 | 0.013 |
|  | GLP-1ra/SGLT-2i | 7,460 (24.0) | 3,428 (10.9) | <0.001 | 0.35 |
| **Clinical Conditions (%)** |  |  |  |  |  |
|  | ACS Unstable Angina | 94 (0.3) | 193 (0.6) | <0.001 | 0.046 |
|  | ACS NSTEMI | 33 (0.1) | 48 (0.2) | 0.132 | 0.013 |
|  | ACS STEMI | 35 (0.1) | 23 (0.1) | 0.137 | 0.013 |
|  | Heart Failure Unspecified | 158 (0.5) | 393 (1.2) | <0.001 | 0.079 |
|  | Heart Failure Systolic | 69 (0.2) | 76 (0.2) | 0.665 | 0.004 |
|  | Heart Failure Diastolic | 29 (0.1) | 55 (0.2) | 0.007 | 0.022 |
|  | Type 1 diabetes mellitus | 194 (0.6) | 393 (1.2) | <0.001 | 0.065 |
|  | Type 2 diabetes mellitus | 2,216 (7.1) | 4,930 (15.7) | <0.001 | 0.271 |
|  | Hypertension | 4,185 (13.4) | 7,932 (25.2) | <0.001 | 0.301 |
|  | Hypercholesterolemia | 1,047 (3.4) | 2,376 (7.5) | <0.001 | 0.185 |
|  | Anxiety | 299 (1.0) | 691 (2.2) | <0.001 | 0.099 |
|  | Depression | 266 (0.9) | 736 (2.3) | <0.001 | 0.119 |
|  | Substance Abuse Disorder | 7 (0.0) | 26 (0.1) | 0.002 | 0.026 |
|  | Eating Disorder | 6 (0.0) | 11 (0.0) | 0.343 | 0.01 |
|  | Sedentary Behavior | 2 (0.0) | 1 (0.0) | 0.992 | 0.005 |
|  | Hyperthyroidism | 31 (0.1) | 93 (0.3) | <0.001 | 0.044 |

**Abbreviations:** IE = Icosapent Ethyl; DHA/EPA = Omega-3-Acid Ethyl Esters; SMD = Standardized Mean Difference; ARB = Angiotensin II Receptor Blocker; ARNI = Angiotensin Receptor-Neprilysin Inhibitor; ACE = Angiotensin-Converting Enzyme Inhibitor; GLP-1 = Glucagon-Like Peptide-1 Receptor Agonist; SGLT-2 = Sodium-Glucose Cotransporter-2 Inhibitor

The PS and exact matching created an analytic cohort having a balanced participant characteristics (SMD < 0.1) between the ICP and DHA/EHA treatment groups with 17,626 participants in each arm. The matched cohort had a median age of 56 (IQR 49, 62) years in both groups, and similar proportion of male (65.8% of ICP vs. 64.3% of DHA/EPA). Beta blockers (38.6% in the ICP group vs. 33.0% in the DHA/EPA group), ACEi (30.7% in the ICP group vs. 32.7% in the DHA/EPA group), ARBs (28.4% in the ICP group vs. 27.6% in the DHA/EPA group), and calcium channel blockers (24.3% in the ICP group vs. 21.5% in the DHA/EPA group) use were well balanced and prevalently utilized in both ICP and DHA/EPA arms. Regarding comorbidities, HTN was present in 18.3% of the ICP group and 16.2% of the DHA/EPA group, while type 2 diabetes mellitus was found in 10.6% of the ICP group and 9.2% of the DHA/EPA group.

The Kaplan-Meier estimates from the matched cohort indicated that the cumulative incidence of AF at 1 year was 1.75 incidence expressed as a % (95% CI: 1.51-2.038) for ICP users and 1.56% (95% CI: 1.33-1.83) for DHA/EPA users. At 2 years, the cumulative incidence was 5.38% (95% CI: 4.782-6.038) and 4.12% (95% CI: 3.63-4.68), respectively. The 2-year HR for AF was 1.242 (95% CI: 1.061 to 1.455) as estimated by the Cox-proportional hazards regression model [Table 2]. The direction and statistical significance of the HR were consistent across a series of multivariable regression analyses, where adjustments were made for each residual potential confounder [Table 3].

**TABLE 2:**
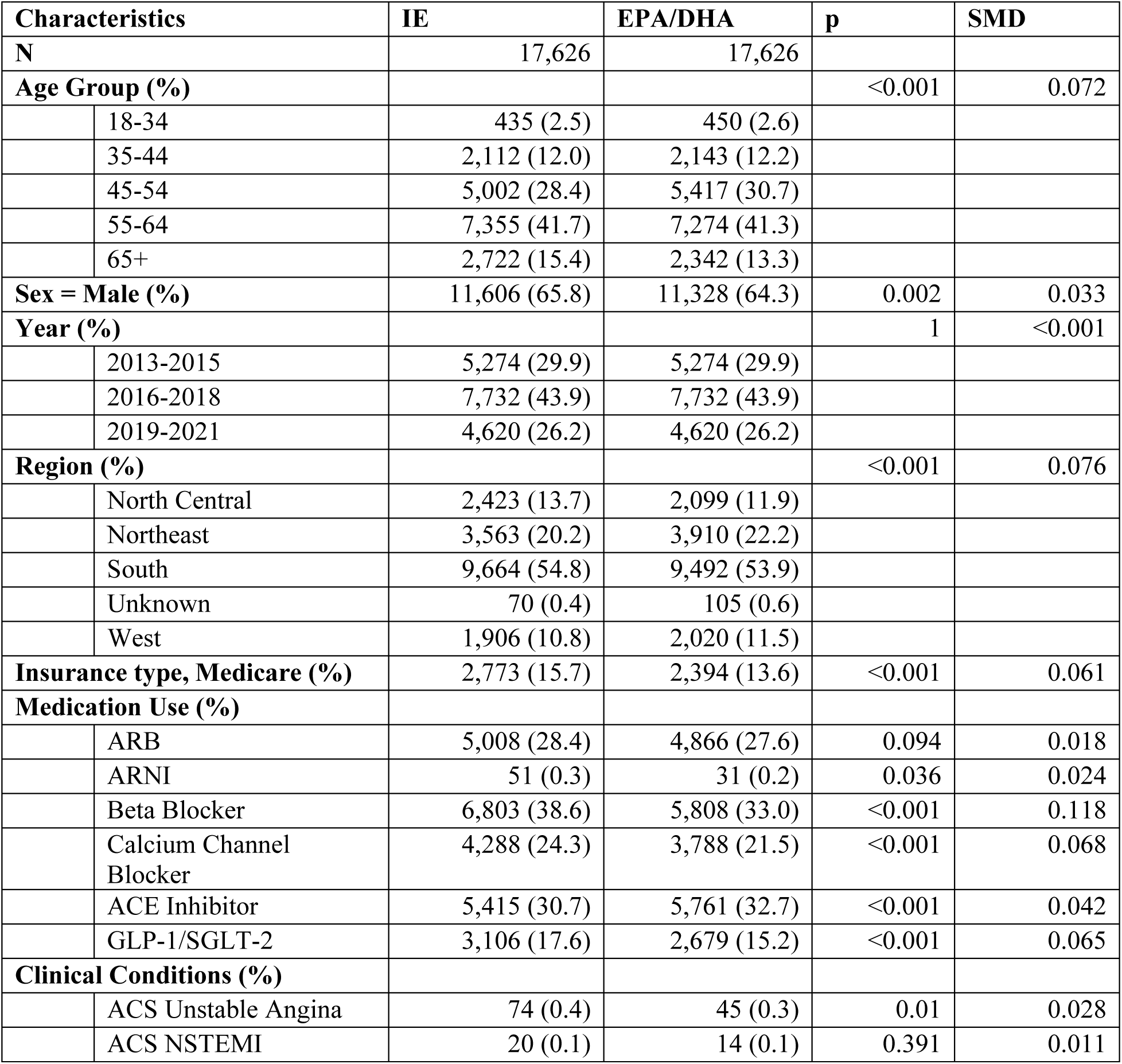
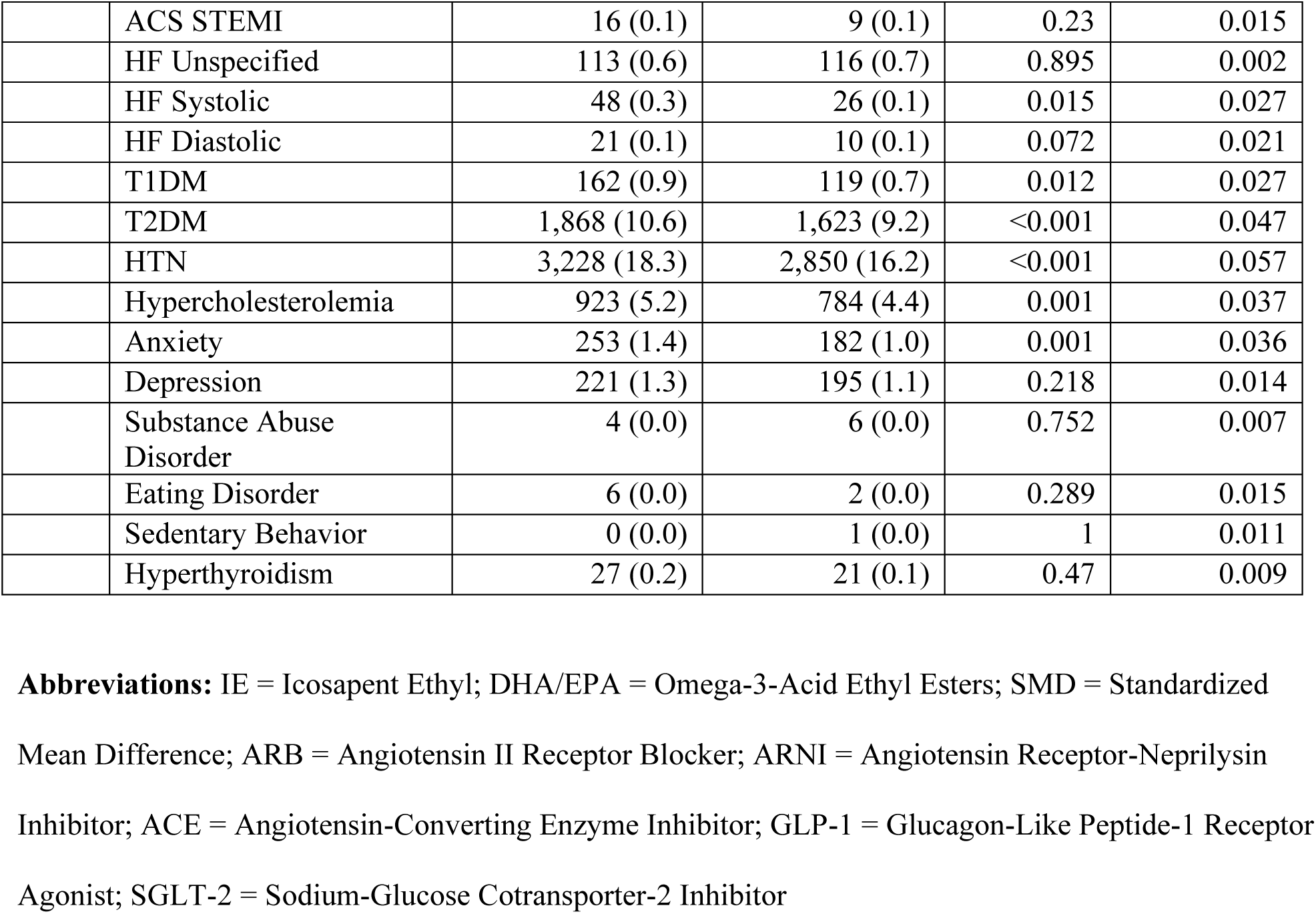
Demographics and clinical characteristics of propensity score matched cohort.

| Characteristics |  | IE | EPA/DHA | p | SMD |
| --- | --- | --- | --- | --- | --- |
| **N** |  | 17,626 | 17,626 |  |  |
| **Age Group (%)** |  |  |  | <0.001 | 0.072 |
|  | 18-34 | 435 (2.5) | 450 (2.6) |  |  |
|  | 35-44 | 2,112 (12.0) | 2,143 (12.2) |  |  |
|  | 45-54 | 5,002 (28.4) | 5,417 (30.7) |  |  |
|  | 55-64 | 7,355 (41.7) | 7,274 (41.3) |  |  |
|  | 65+ | 2,722 (15.4) | 2,342 (13.3) |  |  |
| **Sex = Male (%)** |  | 11,606 (65.8) | 11,328 (64.3) | 0.002 | 0.033 |
| **Year (%)** |  |  |  | 1 | <0.001 |
|  | 2013-2015 | 5,274 (29.9) | 5,274 (29.9) |  |  |
|  | 2016-2018 | 7,732 (43.9) | 7,732 (43.9) |  |  |
|  | 2019-2021 | 4,620 (26.2) | 4,620 (26.2) |  |  |
| **Region (%)** |  |  |  | <0.001 | 0.076 |
|  | North Central | 2,423 (13.7) | 2,099 (11.9) |  |  |
|  | Northeast | 3,563 (20.2) | 3,910 (22.2) |  |  |
|  | South | 9,664 (54.8) | 9,492 (53.9) |  |  |
|  | Unknown | 70 (0.4) | 105 (0.6) |  |  |
|  | West | 1,906 (10.8) | 2,020 (11.5) |  |  |
| **Insurance type, Medicare (%)** |  | 2,773 (15.7) | 2,394 (13.6) | <0.001 | 0.061 |
| **Medication Use (%)** |  |  |  |  |  |
|  | ARB | 5,008 (28.4) | 4,866 (27.6) | 0.094 | 0.018 |
|  | ARNI | 51 (0.3) | 31 (0.2) | 0.036 | 0.024 |
|  | Beta Blocker | 6,803 (38.6) | 5,808 (33.0) | <0.001 | 0.118 |
|  | Calcium Channel Blocker | 4,288 (24.3) | 3,788 (21.5) | <0.001 | 0.068 |
|  | ACE Inhibitor | 5,415 (30.7) | 5,761 (32.7) | <0.001 | 0.042 |
|  | GLP-1/SGLT-2 | 3,106 (17.6) | 2,679 (15.2) | <0.001 | 0.065 |
| **Clinical Conditions (%)** |  |  |  |  |  |
|  | ACS Unstable Angina | 74 (0.4) | 45 (0.3) | 0.01 | 0.028 |
|  | ACS NSTEMI | 20 (0.1) | 14 (0.1) | 0.391 | 0.011 |
|  | ACS STEMI | 16 (0.1) | 9 (0.1) | 0.23 | 0.015 |
|  | HF Unspecified | 113 (0.6) | 116 (0.7) | 0.895 | 0.002 |
|  | HF Systolic | 48 (0.3) | 26 (0.1) | 0.015 | 0.027 |
|  | HF Diastolic | 21 (0.1) | 10 (0.1) | 0.072 | 0.021 |
|  | T1DM | 162 (0.9) | 119 (0.7) | 0.012 | 0.027 |
|  | T2DM | 1,868 (10.6) | 1,623 (9.2) | <0.001 | 0.047 |
|  | HTN | 3,228 (18.3) | 2,850 (16.2) | <0.001 | 0.057 |
|  | Hypercholesterolemia | 923 (5.2) | 784 (4.4) | 0.001 | 0.037 |
|  | Anxiety | 253 (1.4) | 182 (1.0) | 0.001 | 0.036 |
|  | Depression | 221 (1.3) | 195 (1.1) | 0.218 | 0.014 |
|  | Substance Abuse Disorder | 4 (0.0) | 6 (0.0) | 0.752 | 0.007 |
|  | Eating Disorder | 6 (0.0) | 2 (0.0) | 0.289 | 0.015 |
|  | Sedentary Behavior | 0 (0.0) | 1 (0.0) | 1 | 0.011 |
|  | Hyperthyroidism | 27 (0.2) | 21 (0.1) | 0.47 | 0.009 |

**Abbreviations:** IE = Icosapent Ethyl; DHA/EPA = Omega-3-Acid Ethyl Esters; SMD = Standardized Mean Difference; ARB = Angiotensin II Receptor Blocker; ARNI = Angiotensin Receptor-Neprilysin Inhibitor; ACE = Angiotensin-Converting Enzyme Inhibitor; GLP-1 = Glucagon-Like Peptide-1 Receptor Agonist; SGLT-2 = Sodium-Glucose Cotransporter-2 Inhibitor

**Table 3:** Results from Kaplan Meier Estimates and Cox Proportional Hazard Model showing the cumulative incidence and risk of atrial fibrillation among patients treated with ICP and DHA/EPA.

|  | Icosapent Ethyl | Omega-3 Acid Ethyl Ester |
| --- | --- | --- |
| **1year Cumulative Incidence (95% CI)** | 1.75% (1.51-2.038) | 1.56% (1.33-1.83) |
| **2year Cumulative Incidence (95% CI)** | 5.38% (4.782-6.038) | 4.12% (3.63-4.68) |
| **Hazard Ratio (95% CI)- IE vs DHA/EPA** | 1.242 (1.084 to 1.400) |  |

## DISCUSSION

In a large population-based observational cohort study using a United States-based commercial insurance and Medicare supplemental claims database, we observed that ICP significantly increased the cumulative incidence of AF compared to DHA/EPA products, particularly at one year, with the incidence continuing to rise in the second year. This association remained a significant event after adjusting for the increasing prescription volume of ICP over time and controlling for potential confounding factors.

When comparing our results to published randomized clinical trials using ICP or DHA/EPA medications, the REDUCE-IT trial evaluated ICP at a dosage of 4g daily in patients with established CV disease or diabetes and other risk factors, demonstrating a significant reduction in major adverse cardiac event (MACE).^2^ The use of ICP was also associated with higher rate of hospitalization for AF or flutter in the ICP group compared to placebo (5.3% vs. 3.9%, *p*=0.003) and AF requiring hospitalization (3.1% vs 2.1%, *p*=0.004).^2^ In-study hospitalization rates were higher in patients with prior AF, but were substantially lower in patients without prior AF.^4^ In our real-world population, the cumulative AF rate closely mirrors the AF rate from the randomized clinical trial; however, our data suggests that the AF rate in patients without prior AF is 2.5 times higher than reported in the REDUCE-IT study. Our study population consists of a younger, predominantly male, primary prevention cohort compared to the REDUCE-IT study, potentially placing more patients at risk for ICP-associated AF than previously reported. In addition, beyond the trial setting, risk of AF is less aggressively controlled which is captured from our real-world data study.

Focusing on the comparator combination in this trial, the STRENGTH trial studying omega-3 carboxylic acids (combination of DHA/EPA) at 4 g daily in high-risk patients, found no significant CV benefit, but reported an increased risk of AF in the DHA/EPA group compared to placebo (2.2% vs 1.3%, *p*<0.001).^5^ The OMEMI trial, studying 1.8 g of DHA/EPA compared to placebo, in elderly patients with a recent acute myocardial infarction, found no significant reduction in MACE, but observed a higher incidence of AF in the DHA/EPA group that did not reach statistical significance.^6^ In the observational MESA study enrolling individuals without CV disease, higher plasma levels of DHA (but not EPA) were associated with a lower risk of incident AF.^7^ A recent systematic review and meta-analysis of 7 large cardiovascular randomized clinical trials found the use of marine ὠ-3 fatty acid supplementation increased the risk of AF by HR, 1.25 (95% CI: 1.07-1,46) with an increase in HR when stratified by dose.^8^ Thereby, there appears to be contrasting effect of greater CV event efficacy offset by higher risk of AF with increasing omega-3 fatty acid dosing.

Omega-3 fatty acids stabilize the cardiac membrane and protect against arrhythmias, through both direct electrophysiological actions and positive effects on biological processes involved in atrial remodeling.^8^ However, previous reports of alteration in the ventricular action potentials by omega-3 fatty acids through ionic current modification, may potentially promotere-entrant arrhythmias.^9^ Studies have found that mechanically activated nonselective cation channels, PIEZO-1, may mediate the effect of omega-3 fatty acids on AF.^10^ These mechanosensitve channels form homotrimeric dome-shaped structures with three upward curved propeller-like blades and a central pore during assembly.^8, 11^ The blades flatten due to increased mechanical force, which causes the central pore to open, leading to non-selective influx of positively charged ions.^8, 11^ Dietary fatty acids are membrane lipid components that regulate ion channel function.^12–13^ DHA slows channel inactivation of PIEZO-1 while EPA speeds it up.^8^ Therefore, the DHA/EPA ratio might determine the overall effect. A DHA-predominant effect would result in PIEZO1 activation and increased influx of calcium and other cations, potentially prolonging the action potential leading to delayed after-depolarizations resulting in AF.^8^ In addition, atrial fibroblasts from AF patients have higher PIEZO-1 expression levels and activity than those from patients with sinus rhythm, indicating that PIEZO-1 may promote atrial structural remodeling.^11^ Other reasons for DHA/EPA causing a pro-arrhythmic environment may result from changes in voltage-gated ion channels, ionic pumps, cell surface receptors, and extracellular matrix-cytoskeletal interactions.^8^ Further mechanistic basic and dose-dependent clinical trials are required to understand the processes by which omega-3 fatty acid supplementation increases the AF risk and alters the electrophysiological characteristics of the myocardium.

### Limitations

Our study has several limitations that should be considered. Firstly, we were unable to capture the dispensing of over the counter (OTC) fish oils. However, it is unlikely that patients would concurrently take ICP or DHA/EPA together with OTC fish oils. Even with a small chance of concurrent use of prescription ICP or DHA/EPA with OTC fish oils, the pattern of the OTC medication or dietary supplement use would not be differential between the ICP and DHA/EPA. Therefore, the measure of association calculated in HR will not be or minimally influenced by the baseline or concurrent OCT fish oil use. We also did not account for changes in statin dosage or the alteration of other medications after the follow-up period began. While we believe that our matching strategy relieved the concern on the difference in the patterns of the use of the other agents or statin dose alterations between the ICP and DHA/EPA arms, future research could benefit from clinical trials that more rigorously control these variables. We did not follow-up on participant AF outcomes in this study. Understanding of the consequences of AF, including self-termination post discontinuation of ICP or DHA/EPA, need for therapeutic interventions, potential for development of paroxysmal or permanent AF, and healthcare costs are important to understand for patients considering taking these agents or supplements. Finally, yet importantly, any interpretation of our findings should be accompanied by the general limitations of retrospective observational research, such as coding errors, limited generalizability, or unobserved confounders, while the influence of such limitations would be nominal on our association measures.

## CONCLUSIONS

In adult naïve AF-naive patients, ICP, compared to DHA/EPA supplements taken with statins, was associated with a higher risk of developing AF.

## Data Availability

Data is available on request from the authors.

## Author Contributions

All authors fully contributed to the content of this manuscript, including meeting the four criteria of the Internal Committee of Medical Journal Editors. All authors had full access to all the data in the study and took full responsibility for integrity of the work and accuracy of the data analysis, from inception to published article.

## Analytic Personnel

Jyotirmoy Sarker, M.Pharm., M.B.A.., Michael Kim, Pharm.D., Mark A. Munger, Pharm.D., Kibum Kim, Ph.D.

## ABREVIATIONS

ACEi: Angiotensin-converting enzyme inhibitor
ARB: Angiotensin receptor blocker
ARNI: Angiotensin Receptor-Neprilysin Inhibitor
AF: Atrial Fibrillation
BB: ßeta-adrenergic blocker
DHA/EPA: Eicosapentaenoic and doxosahexaenoic acid
EPA: Eicosapentaenoic acid
GLP-1ra: Glucoagon-like-peptide-1 receptor agonists.
HR: Hazard Ratio
ICP: Icopasent Ethyl
MACE: Major Adverse Cardiac Event
PIEZO: A mechanosensitive ion protein
PS: Propensity Score
SGLT2i: Sodium-glucose cotransporter-2 inhibitors
SPL: Spironolactone
SMD: Standard Mean Difference

